# Quantifying Inaccuracies in Modeling COVID-19 Pandemic within a Continuous Time Picture

**DOI:** 10.1101/2020.09.05.20188755

**Authors:** Ioan Bâldea

## Abstract

Typically, mathematical simulation studies on COVID-19 pandemic forecasting are based on deterministic differential equations which assume that both the number (*n*) of individuals in various epidemiological classes and the time (*t*) on which they depend are quantities that vary continuous. This picture contrasts with the discrete representation of *n* and *t* underlying the real epidemiological data reported in terms daily numbers of infection cases, for which a description based on finite difference equations would be more adequate. Adopting a logistic growth framework, in this paper we present a quantitative analysis of the errors introduced by the continuous time description. This analysis reveals that, although the height of the epidemiological curve maximum is essentially unaffected, the position 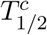 obtained within the continuous time representation is systematically shifted backwards in time with respect to the position 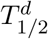 predicted within the discrete time representation. Rather counterintuitively, the magnitude of this temporal shift 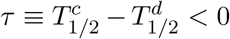 is basically insensitive to changes in infection rate *κ*. For a broad range of *κ* values deduced from COVID-19 data at extreme situations (exponential growth in time and complete lockdown), we found a rather robust estimate *τ* ≃ −2.65 day^−1^. Being obtained without any particular assumption, the present mathematical results apply to logistic growth in general without any limitation to a specific real system.

## 1 Introduction

Since the COVID-19 infectious disease caused by the severe acute respiratory syndrome coronavirus 2 (SARS-CoV-2) broke out in Wuhan City, the capital of Hubei province of China,^1,2^ on December 2019 and WHO declared it a pandemic,^3,4^ collaborative work was conducted worldwide by research institutions and pharma industry aiming at developing antiviral medication and vaccine in the global battle against this unprecedented dangerous threat to planet’s life. Along with such effort whose concrete results are still to come, mathematical simulation can significantly assist governments in adopting the most adequate solutions to suppress infection spread and minimize medical burden as well as dramatic economic and social consequences.

Most population dynamics studies on COVID-19 pandemic as well as on other epidemics carried out in the past adopted a mathematical framework based on a continuous time picture. This is particularly comfortable because it enables to derive the temporal evolution of the populations of various epidemiological compartments by solving a set ordinary differential equations. Still, a more realistic approach should aim at quantitatively predicting or at least reproducing the new infection cases, and these are reported daily. Therefore, a discrete time representation appears to be more adequate. From this perspective, quantifying the errors arising from the simplified continuous time assumption is a significant question that deserves consideration. However, to the best of author’s knowledge, this issue was not addressed in previous COVID-19 studies. For this reason, it represents the focus of the present paper.

## 2 The Logistic Model

Evolution in time of epidemics was extensively investigated in the literature, most often within the celebrated SIR model or many of its extensions.^5–10^ An important problem with such approaches is the (too) large number of input parameters they need in order to make reliable predictions, and their values are difficult to validate before disease end.^11^

For this reason, we will adopt here the simpler logistic growth model.^12–15^ Along with numerous applications in various other fields,^16–21^ this model turned out to be also useful in previous studies on epidemics,^22–25^ also including COVID-19 pandemic.^26–28^ As compared to various SIR-based approaches, the logistic growth model possesses a great advantage: it can be directly validated against ongoing epidemiological reports.^28^

In the continuous time description, logistic growth in time of an infected population obeys a first-order differential equation

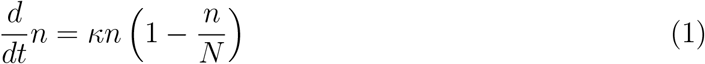

Here, *κ* stands for the intrinsic infection rate, which characterizes the unlimited exponential (Malthus-type) population increase occurring in an infinite environment. In real situations, exponential growth is suppressed because the effective infection rate 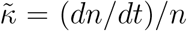 gradually decreases as the number of infections increases. The parenthesis entering the RHS of equation (1) resembles the Pauli blocking factor passionately discussed in charge transport in Fermi systems.^29–33^ It represents the simplest possible way to model a population dependent infection rate 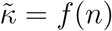, namely to assume that it linearly decreases as *n* increases and eventually levels off at the maximum value *N*, which defines the so-called carrying capacity.

Epidemiologists refer to *n* = *n*(*t*) as the total (cumulative) number of cases and to 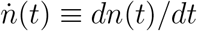 as the daily number of (new) infections. The latter term stems from discrete time counterpart of the temporal derivative

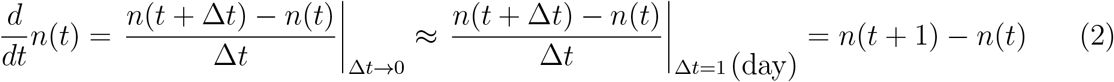

By replacing the derivative entering the LHS of equation (1) by its finite difference counterpart of the RHS of equation (2) we arrive at the discrete time formulation of logistic growth

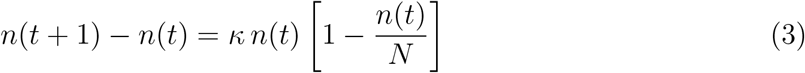

The advantage of the continuous time description of logistic growth is that equation (1) can be integrated out in closed analytical form with the well known results

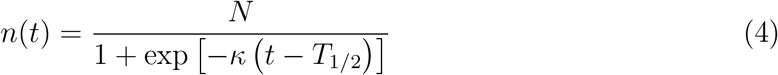

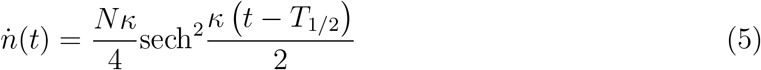

Notice that, whether in continuous or discrete time, in addition to the two model parameters *κ* and *N*, an “initial” condition is needed to determine the time evolution *n* = *n*(*t*) via equations (1) or (3), respectively. Most commonly this is imposed by

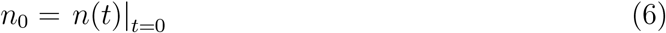

In continuous time description this is often recast in terms of the half-time defined by 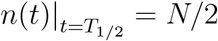 which yields

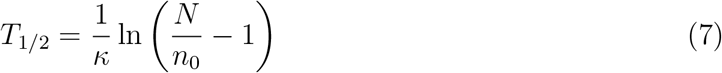

## 3 Results and Discussion

Inspection of equations (1) or (3) reveals that results for the normalized number of cumulative cases *n*(*t*)/*N* and daily cases (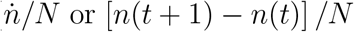, respectively) as a function of the reduced time *t/T*_1/2_ can be depicted as curves depending on a single parameter, namely *κ*; the value *n*_0_/*N* merely serves to appropriately set the time origin. To be specific, guided by our recent study,^28^ in all the figures presented below we used *n*_0_/*N* ≈ 1/1500. However, as just said, this particular choice does by no means affect the generality of the results reported below.

To start our analysis, we chose a value of infection rate *κ* = 0.14 day^−1^, which turned out to describe a regime of moderate restrictions during Slovenia COVID-19 epidemic.^28^ Results for the cumulative and daily number of cases obtained within the continuous time and discrete time using the value *κ* = 0.14 day^−1^ can be compared in Figs. 1b and 2b, respectively. These figures reveal a certain backward shift of the continuous time curves with respect to the discrete time curves.

**Figure 1:**
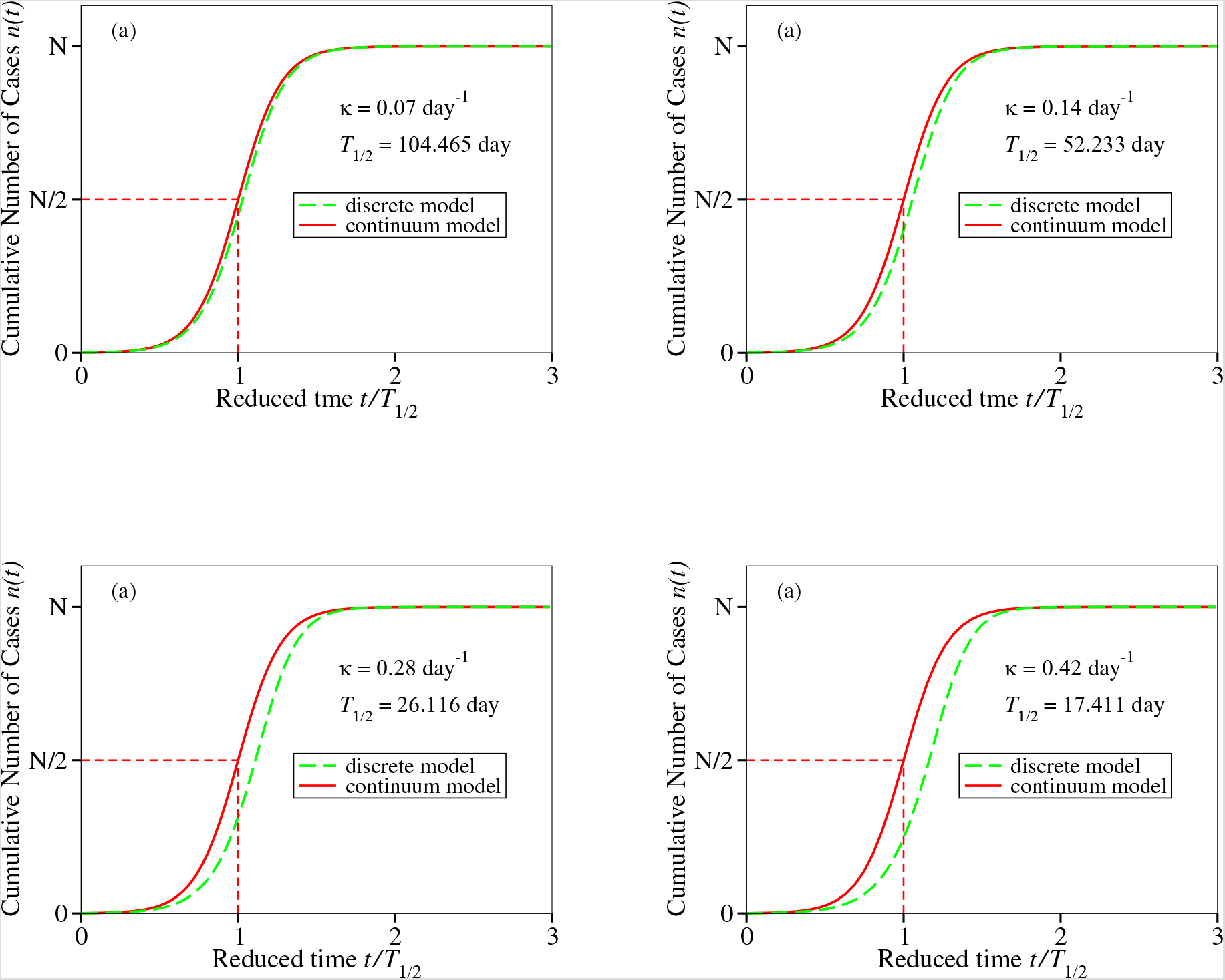
Cumulative number of infection cases described by means of logistic growth models employing continuous and (“exact”) discrete time representation (equations (4) and (2), respectively) for several typical values of the infection rate *κ* indicated in the inset. Time on the *x*-axis is expressed in units of the half-time *T*_1/2_.

**Figure 2:**
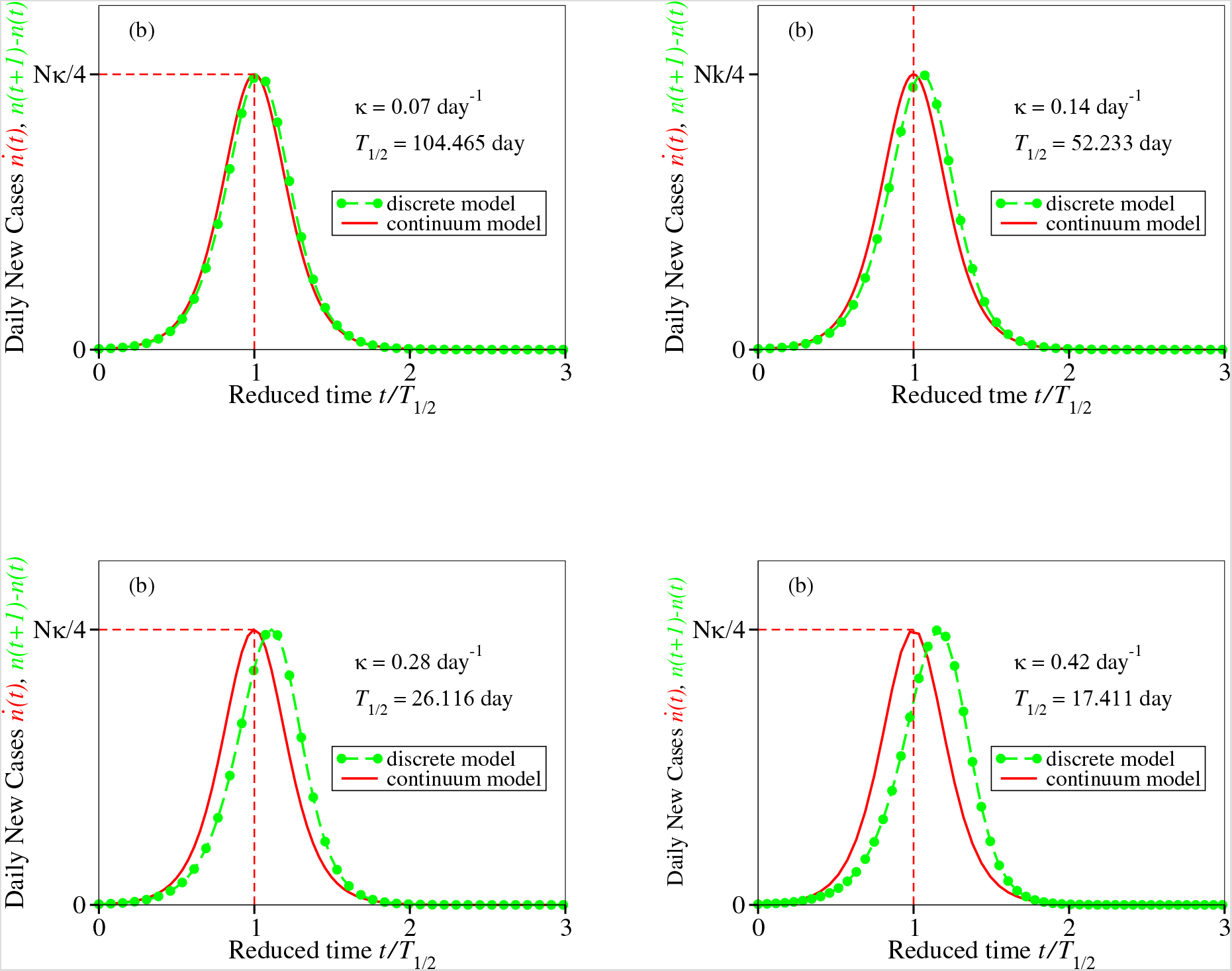
Epidemiological curves depicting daily numbers of new infections described by means of logistic growth models employing continuous and (“exact”) discrete time representation (equations (5) and 2), respectively) for several typical values of the infection rate *κ* indicated in the inset. Time on the *x*-axis is expressed in units of the half-time *T*_1/2_.

Based on naive intuition it seems reasonably to expect that situations corresponding slower temporal variations (smaller infection rates *κ*) can more appropriately be described quantitatively within a continuous time representation than those characterized by faster variations (larger *κ* values). To check whether this is the case or not, we monitored how results change by increasing *κ*. Results for the larger value *κ* = 0.28 day^−1^ are depicted in Figs. 1c and 2c. They show backward shifts larger than for *κ* = 0.14 day^−1^. Further increasing *κ* to the value *κ* = 0.42 day^−1^ (Figs. 1d and 2d) confirms this trend. On the contrary, the curves computed for the smaller value *κ* = 0.07 day^−1^ (Figs. 1a and 2a) reveal that the backward shift is smaller. So far, so good; the intuitive expectation is supported by these calculations. The curves based on continuous time exhibits a systematic shift backwards in time whose magnitude varies monotonically with *κ*. Noteworthily, the behavior displayed in Figs. 1 and 2 refers to quantities plotted against the reduced time *t/T*_1/2_.

Surprise arises when examining the curves plotted with the absolute time *t* on the abscissa rather than the reduced time *t/T*_1/2_. The difference visible in Figs. 1 and 2 is not so clear when using *t* instead of *t/T*_1/2_ on the x axis (see Fig. 3a). However, it becomes observable when the curves are shifted by *T*_1/2_, that is, are plotted versus *t* − *T*_1/2_ (see Fig. 3b) Still, the κ-dependence of the peak heights (they are proportional to *κ*, see equation (5)) makes observation somewhat difficult.

**Figure 3:**
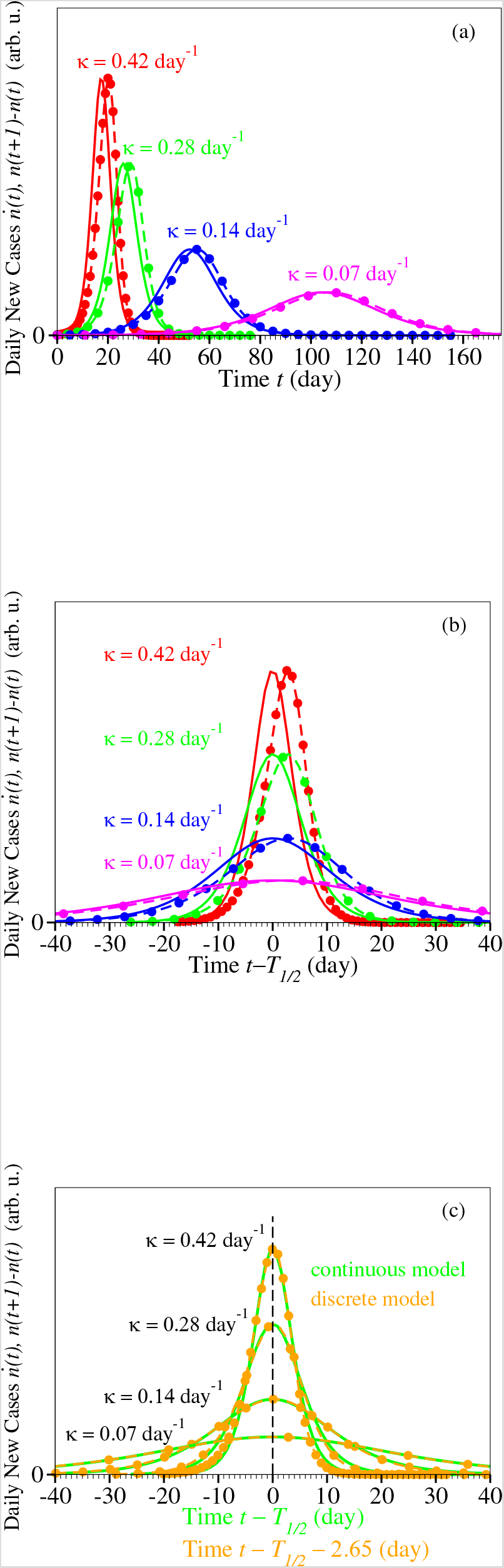
Epidemiological curve depicting daily numbers of new infections described by means of logistic growth models employing continuous and (“exact”) discrete time representation for several typical values of the infection rate *κ* indicated in the inset.

Finally, the point we want to make is obvious in Fig. 3c. For a very broad range of κ-values, the maximum of the epidemiological curves deduced within the continuous time description is shifted backward in time by the *κ*-independent amount *τ* ≃ −2.65 day while its height (= maximum number of daily new cases) cannot be distinguished from that predicted within the (realistic) discrete time description. Concerning the *κ*-independence of *τ*, one should note that that the above value *κ* = 0.42 day^−1^ corresponds to a regime wherein infections were found to growth exponentially in time while *κ* = 0.07 day^−1^ is almost half the *κ*-value at complete lockdown.^28^

To conclude, the above finding contradicts the expectation based on naive intuition that faster population dynamics is poor(er) describable in continuous time, or, put differently, that slower infections are better described within a continuous time approach.

## 4 A Technical Remark

In arriving at equation (2), we chose the forward (finite) difference *n*(*t*+1)−*n*(*t*) to establish the correspondence between the discrete time and continuous time versions of logistic growth. This procedure is just in the vein underlying the logistic growth philosophy. The LHS of equation (1) is nothing but the number of new daily cases. Via equation (1), this number is assumed to be proportional to the number of total of *existing* infections *n*(*t*) as long as the latter are few. Otherwise, the gradual increase in the fraction of the *already* infected individuals *n*(*t*)/*N* reduces the probability for further infection.

Purely mathematically speaking, as alternatives to the forward difference (*dn*(*t*)/*dt* → *n*(*t* + 1) − *n*(*t*)), we could have chosen the backward finite difference (*dn*(*t*)/*dt* → *n*(*t*) − *n*(*t* − 1)) or the symmetric finite difference (*dn*(*t*)/*dt* → [*n*(*t* + 1) − *n*(*t* − 1)] /2).

The backward difference leads to

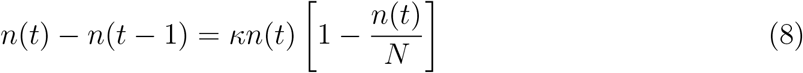

Although difference in numerical results obtained by employing equation (8) and equation (2) are not notable, equation (8) does complain neither with the idea of logistic growth (see above) nor to the causality principle.

Leading to

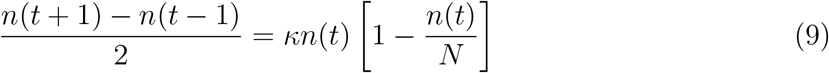

the symmetric finite difference (which is a better choice for calculating numerical derivatives in other cases) is even more problematic here. In addition to the drawback noted in connection with equation (8), unrealistically, the symmetric difference version of equation (9) does not allow to uniquely predict time evolution *n*(*t*) merely based on a single initial condition (*n*(0) = *n*_0_); equation (9) is a relationship between populations at three consecutive days: *n*(*t* + 1), *n*(*t*), and *n*(*t* − 1).

## 5 Conclusion

Based on the present results obtained within a logistic growth model, we conclude that the most notable inaccuracy brought about by the continuous time description of epidemics — which is mathematically more convenient than the discrete time description — is the prediction that the maximum number 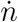 of daily infections occurs earlier in time: 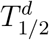 → 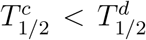. This backward shift in time 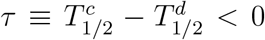 (which does not affect the height of the epi peak) amounts to *τ* ≈ −2.65 day, a robust estimate, which was found to be independent of the infection rate *κ* ranging from large values deduced for situations where COVID-19 infections growth exponentially in time to small values obtained at complete lockdown.^28^ This finding contradicts naive intuition expecting that faster infections (larger *κ*) are poorer described within continuous time approaches than slower infections (smaller *κ*).

To end, let us emphasize that, although conducted in conjunction with the actual unprecedented crisis caused by COVID-19 pandemic, the present study reports results applying to logistic growth in general. No specific reference to a special real system was needed in the mathematical derivation presented above.

## Data Availability

All relevant data have been included in the manuscript.

## Reference

1. http://www.who.int/csr/don/12-january-2020-novel-coronavirus-china/en/, Novel Coronavirus — China. World Health Organization (WHO). Retrieved 9 April 2020.

2. Huang, C.; Huang, Y.; Li, X.; Ren, L.; Zhao, J.; Hu, Y.; Zhang, L.; Fan, G.; Xu, J.; Gu, X. et al. Clinical features of patients infected with 2019 novel coronavirus in Wuhan, China. Lancet 2020, 395, 497–506.

3. http://www.who.int/dg/speeches/detail/who-director-general-s-opening-remarks-at-the-media-briefing-on-covid-19---11-march-2020 “WHO Director-General’s opening remarks at the media briefing on COVID-1911 March 2020”. World Health Organization. 11 March 2020. Retrieved 11 March 2020.

4. Cucinotta, D.; Vanelli, M. WHO Declares COVID-19 a Pandemic. Acta Bio Medica Atenei Parmensis 2020, 91, 157–160.

5. Kermack, W. O.; McKendrick, A. G. Contributions to the mathematical theory of epidemics. I. Proc. Roy. Soc. 1927, 115A, 700–721, reprinted in Bull. Math. Biol. 53, 3355 (1991). https://doi.org/10.1007/BF02464423.

6. Kermack, W. O.; McKendrick, A. G. Contributions to the mathematical theory of epidemicsII. The problem of endemicity. Proc. Roy. Soc. 1932, 138A, 55–83, reprinted in Bull. Math. Biol. 53, 5787 (1991). https://doi.org/10.1007/BF02464424.

7. Kermack, W. O.; McKendrick, A. G. Contributions to the mathematical theory of epidemics. III. Further studies of the problem of endemicity. Proc. Roy. Soc. 1933, 141A, 94–122, reprinted in Bull. Math. Biol. 53, 89118 (1991). https://doi.org/10.1007/BF02464425.

8. Bailey, N. T. J. The Mathematical Theory of Infectious Diseases and Its Applications; Charles Griffin & Company Ltd, 5a Crendon Street, High Wycombe, Bucks HP13 6LE., 1975.

9. Hethcote, H. W. A Thousand and One Epidemic Models. Frontiers in Mathematical Biology. Berlin, Heidelberg, 1994; pp 504–515.

10. Hethcote, H. W. The Mathematics of Infectious Diseases. SIAM Review 2000, 42, 599–653.

11. Fong, S. J.; Li, G.; Dey, N.; Crespo, R. G.; Herrera-Viedma, E. Finding an Accurate Early Forecasting Model from Small Dataset: A Case of 2019-nCoV Novel Coronavirus Outbreak. International Journal of Interactive Multimedia and Artificial Intelligence 2020 6, 132–140.

12. Verhulst, P.-F. Notice sur la loi que la population poursuit dans son accroissement. Correspondance Mathématique et Physique 1838, 10, 113–121.

13. Verhulst, P.-F. Recherches mathémathiques sur la loi d’accroissement de la population. Nouveaux Mémoires de l’Academie Royale des Sciences et Belles-Lettres de Bruxelles 1845, 18, 8.

14. Quetelet, L. A. J. Du Système Social et des Lois qui le Régissent; Guillaumin, 1848.

15. Ostwald, W. Studien zur chemischen Dynamik; Erste Abhandlung: Die Einwirkung der Suren auf Acetamid. Journal für Praktische Chemie 1883, 27, 1–39.

16. McKendrick, A. G.; Pai, M. K. XLV. The Rate of Multiplication of Micro-organisms: A Mathematical Study. Proceedings of the Royal Society of Edinburgh 1912, 31, 649–653.

17. Lloyd, P. American, German and British antecedents to Pearl and Reed’s logistic curve. Population Studies 1967, 21, 99–108.

18. Cramer, J. The early origins of the logit model. Studies in History and Philosophy of Science Part C: Studies in History and Philosophy of Biological and Biomedical Sciences 2004, 35, 613–626.

19. Vandermeer, J. How Populations Grow: The Exponential and Logistic Equations. Nature Education Knowledge 2010, 3, 15.

20. Bâldea, I. Floppy Molecules as Candidates for Achieving Optoelectronic Molecular Devices without Skeletal Rearrangement or Bond Breaking. Phys. Chem. Chem. Phys. 2017, 19, 30842–30851.

21. Bâldea, I. A sui generis electrode-driven spatial confinement effect responsible for strong twisting enhancement of floppy molecules in closely packed self-assembled monolayers. Phys. Chem. Chem. Phys. 2018, 20, 23492–23499.

22. Mansfield, E.; Hensley, C. The Logistic Process: Tables of the Stochastic Epidemic Curve and Applications. Journal of the Royal Statistical Society: Series B (Methodological) 1960, 22, 332–337.

23. Waggoner, P. E.; Aylor, D. E. Epidemiology: A Science of Patterns. Annual Review of Phytopathology 2000, 38, 71–94, PMID: 11701837.

24. Koopman, J. Modeling Infection Transmission. Annual Review of Public Health 2004, 25, 303–326, PMID: 15015922.

25. Bangert, M.; Molyneux, D. H.; Lindsay, S. W.; Fitzpatrick, C.; Engels, D. The crosscutting contribution of the end of neglected tropical diseases to the sustainable development goals. Infect Dis. Poverty 2017, 6, 73.

26. Hermanowicz, S. W. Simple Model for Covid-19 Epidemics - Back-casting in China and Forecasting in the US. medRxiv 2020, DOI 10.1101/2020.03.31.20049486.

27. Bâldea, I. Suppression of Groups Intermingling as Appealing Option For Flattening and Delaying the Epidemiologic Curve While Allowing Economic and Social Life at Bearable Level During COVID-19 Pandemic. medRxiv 2020, DOI 0.1101/2020.05.25.20112938.

28. Bâldea, I. What Can We Learn from the Time Evolution of COVID-19 Epidemic in Slovenia? medRxiv 2020, DOI 10.1101/2020.05.25.20112938.

29. Datta, S. Exclusion principle and the Landauer-Büttiker formalism. Phys. Rev. B 1992, 45, 1347–1362.

30. Sols, F. Scattering, dissipation, and transport in mesoscopic systems. Ann. Phys. (NY) 1992, 214, 386–438.

31. Bönig, L.; Schönhammer, K. Pauli principle in the theory of nonlinear electronic transport. Phys. Rev. B 1993, 47, 9203–9207.

32. Datta, S. Electronic Transport in Mesoscopic Systems; Cambridge Univ. Press: Cambridge, 1997.

33. Wagner, M. Probing Pauli Blocking Factors in Quantum Pumps with Broken Time-Reversal Symmetry. Phys. Rev. Lett. 2000, 85, 174–177.

